# The Cost-Effectiveness of Group B Streptococcus Screening Strategies in Pregnant Women for the Prevention of Newborn Early-onset Group B Streptococcus : A Systematic Review

**DOI:** 10.1101/2024.08.25.24312541

**Authors:** CL Allen, E Naznin, T J R Panneflek, T Lavin, M E Hoque

**Affiliations:** Department of Medicine, Nursing and Health Sciences, Monash University, Melbourne, Australia; International Union Against Tuberculosis and Lung Disease, Asia Pacific Region, Australia; Division of Neonatology, Department of Paediatrics, Willem-Alexander Children’s Hospital, Leiden University Medical Centre, Leiden, the Netherlands; UNDP-UNFPA-UNICEF-WHO-World Bank Special Programme of Research, Development and Research Training in Human Reproduction (HRP), Department of Sexual and Reproductive Health and Research, World Health Organization, Geneva, Switzerland; School of Population and Global Health, University of Western Australia, Perth, Australia

## Abstract

**Background:** Early-onset Group B Streptococcus (EOGBS) infection is one of the most prevalent neonatal infections globally, contributing to significant infant morbidity and mortality by inducing life threatening sequelae such as sepsis, meningitis and pneumonia. EOGBS infection occurs within 7 days of birth following vertical transmission of the bacteria from a colonised pregnant woman to her infant. Current strategies aimed at preventing EOGBS focus on the administration of intrapartum antibiotic prophylaxis (IAP). There is no universally agreed upon strategy for how to best identify which pregnant women should receive IAP. Currently there are four main strategies employed by health systems: 1) risk -based approach where women are assessed for risk factors for newborn EOGBS and IAP is administered to women who have at least one risk factor; 2) universal screening where all women are screened antenatally for GBS colonisation and are given IAP upon testing positive; 3) a combination of a risk-based approach and universal screening, and 4) no strategy for screening strategy with IAP administered on a case-by-case basis. Despite evidence suggesting that a universal screening strategy may be most efficacious in reducing EOGBS incidence, each screening strategy carries with it different costs and economic burdens, depending on the setting. Therefore, recommendations as to which screening strategy is most suitable must be made in the context of both sound clinical and economic evidence.

**Methods:** This review synthesised and compared economic evaluations of maternal GBS screening strategies. A systematic search for evidence relating to GBS screening strategies was performed in the databases MEDLINE, Embase and Web of Science. Studies were included if they reported on a strategy to assess women for IAP administration and the outcomes of interest. This paper presents the findings of economic evaluations identified by this search. The economic findings of each study were compared and synthesised narratively due to significant heterogeneity among included studies preventing meta-analysis.

**Results:** A total of 18 studies were identified for inclusion in this review. These studies, all from high-income countries, cumulatively made 58 comparisons of GBS screening strategies and cost-effectiveness analyses. Studies either compared any type of screening to no screening strategy (Universal screening vs no screening; risk-based approach vs no screening; combined screening vs no screening) or compared different screening strategies to each other. The implementation of any screening strategy was found to be cost-effective compared to none at all depending on the setting (one instance using universal screening, two using risk-factor approach and four using a combined strategy). On multiple occasions, cost-effectiveness varied significantly depending on the prevalence of maternal GBS colonisation.

**Discussion:** This review demonstrated that in several instances the implementation of any GBS screening strategy was cost-effective compared to no strategy at all. Greater evidence is required to determine which type of screening strategy is most cost-effective, particularly in lower resource settings. The variability of cost-effectiveness by prevalence of maternal GBS colonisation indicates that a strategy’s economic viability is likely context specific and should be considered before the implementation of any screening strategy.

## Introduction

Early-onset Group B Streptococcus (EOGBS) refers to an infection caused by the bacterium Streptococcus agalactiae (Group B streptococcus or GBS) that occurs in newborn infants within the first week of life; typically within the first 24 to 48 hours after birth . EOGBS infections can be caused by vertical transmission of GBS colonised pregnant women (vagina or rectum) to an infant. The global prevalence of rectovaginal GBS colonisation among pregnant women is estimated to be between 11% and 30% at the time of birth, dependent on geographical location (1). Transmission to the baby during birth occurs in around 70% of instances where the mother has GBS colonisation (2). EOGBS infections can manifest in various forms in the newborn, including septicaemia blood infection), pneumonia (lung infection) and meningitis (infection of the membranes covering the brain and spinal cord) (3, 4). These infections can lead to serious health complications such as encephalopathy (brain dysfunction), and, in severe cases, can be life-threatening for the newborn.

EOGBS burden has emerged as a challenge facing global health systems with large disparities in prevalence between regions. In 2022 there were 231,800 EOGBS cases globally with the majority of these cases occurring in Sub-Saharan Africa and Central and Southern Asia (5). The loss of life associated with EOGBS is considerable, as an estimated 58,300 deaths were thought to have resulted from the disease in 2022 alone. Similar to prevalence, the largest mortality burden of EOGBS is borne by health systems in Sub-Saharan Africa and Central and Southern Asia (5). The impact of early-onset Group B Streptococcal (EOGBS) infection in newborns extends far beyond the immediate health challenges. For families and communities, the emotional toll of caring for a newborn with EOGBS, especially when the infection results in death, is profound. The financial burden of providing specialized care for these newborns can place a considerable strain on healthcare systems, particularly in under-resourced regions where the prevalence of EOGBS is often among the highest. The collective cost to the family, community, and society at large is substantial, underscoring the need for more support and resources to manage this serious condition effectively. Preventing the transmission of the GBS bacteria between mother and baby and subsequently reducing rates of EOGBS is therefore vital to alleviating the high burden (5).

To prevent the transmission of GBS bacteria from the mother to newborn, pregnant women can be given intravenous antibiotics during labour ((intrapartum antibiotic prophylaxis (IAP)) (6). IAP has been found to reduce the incidence of EOGBS by approximately 80% and has significantly reduced EOGBS related mortality (7, 8). However, due to concerns about antimicrobial resistance, rising non-GBS early-onset infection and antibiotic exposure, administering antibiotics to all women in labour is not feasible nor ethical (9). Identifying which women would benefit from IAP is a challenge, though various strategies exist. Firstly, universal screening strategies, test pregnant women for GBS colonization using vaginal-recto swabs during antenatal care visits (healthcare visits during pregnancy). Universal strategies require the sample to be taken with microbiological testing in a laboratory to detect for GBS colonisation. Pregnant women who are found to have GBS colonisation are subsequently given IAP at the time of labour (10). Universal strategies require significant resource investment in establishing a programme that routinely screens all pregnant women with access to laboratories to process the samples.

Secondly, a risk-based approach may be used where pregnant women are assessed for risk-factors for neonatal EOGBS around the time of birth with IAP administered to women who have one or more risk factor. Risk factors can include i) a previous infant affected by (EO)GBS infection, ii) GBS bacteriuria during current pregnancy, iii) intrapartum maternal fever, iv) preterm labour, or v) prolonged rupture of membranes (10). Abiding by a risk-based approach means that IAP is administered to every pregnant woman who presents with one or more risk-factor, however the risk-factors used in each setting can vary (10).

Thirdly, in some settings a combination of risk-based approach and universal strategies are used. For example, one combined strategy is that all pregnant women have universal microbiological testing during antenatal care, and IAP is administered to pregnant women positive for GBS colonization as well as at least one risk-factor.

Finally, in some settings IAP is provided on an individual basis without consistent rules or criteria (11). Previous systematic reviews and meta-analyses of observational evidence demonstrate that any screening strategy as compared to no strategy may be associated with reduced incidence of EOGBS; and that universal screening may be associated with a reduced incidence of EOGBS as compared to a risk-based strategy (12).

Trade-offs between the benefits to newborns for implementing a screening programme and the costs and resource investment needed in coordinating such programmes, staff training, investment in establishing laboratories needs careful consideration (13). There has been limited research mapping the cost-effectiveness of different GBS screening strategies. A 2022 scoping review of economic evaluations of maternal health interventions by Eddy et al. (14) identified 11 studies that were economic evaluations or contained economic evaluations of interventions relating to GBS. To our knowledge, there is no existing systematic review comparing economic evaluations of different GBS screening strategies.

This systematic review aims to identify and synthesise economic evaluations of GBS screening strategies. This includes studies that have assessed the cost-effectiveness, cost-benefit, or cost-utility of GBS screening strategies including: risk-based approaches, universal strategies and combined strategies as compared to each other or no screening strategy. This review did not cover cost-effectiveness analysis for GBS vaccines, rapid intrapartum PCR testing, or routine IAP to all women.

## Methods

The objective of this study was to conduct a systematic review to assess the evidence and knowledge gaps in the published literature on the cost-effectiveness of Group B Streptococcus screening strategies.

### Protocol and registration

The Preferred Reporting Items for Systematic Reviews and Meta-Analyses (PRISMA) guidelines and Cochrane Handbook for Systematic Reviews were used to conduct and report this systematic review (Suppl. Table 1) (15, 16). The full protocol which encompassed a systematic review on GBS screening strategies for a number of maternal and newborn outcomes was registered in PROSPERO, an international prospective register of systematic reviews, with ID CRD42023411806 (17). This current manuscript refers to the cost-effectiveness component of the full protocol.

#### Inclusion criteria

Studies were included when they provided data on economic evaluations comparing a screening strategy with no screening strategy, or another screening strategy. Economic studies eligible for inclusion in this review include cost-effectiveness analyses (CEA), cost-benefit analyses (CBA), cost-utility analyses (CUA) and cost-consequence analyses (CCA). Studies that were not stand-alone economic evaluations but that included an economic evaluation as part of a larger study such as randomised controlled trials, non-randomised intervention studies, or observational studies were eligible to be included. Studies were eligible to be included if they reported discrete health outcomes e.g. lives saved, cases of EOGBS prevented or composite outcomes e.g., disability adjusted life years (DALY) or quality adjusted life years (QALY). We considered studies that used primary or secondary data.

#### Exclusion criteria

Studies were excluded if they did not fulfil the criteria to be deemed full economic evaluations (e.g., partial economic evaluations). Editorials, protocols, poster presentations, letters and review articles were also excluded.

#### Search strategy

The search was carried out on across the MEDLINE, Embase and Web of Science databases 18 May 2023 with an updated search in May 2024. Details of this search, including an example of the search strategy are presented in Suppl. Table 2 and the PRISMA flowchart are included (Figure 1). A snowball method through hand-searching of grey literature and checking the reference list of included publications was conducted in parallel to ensure completeness.

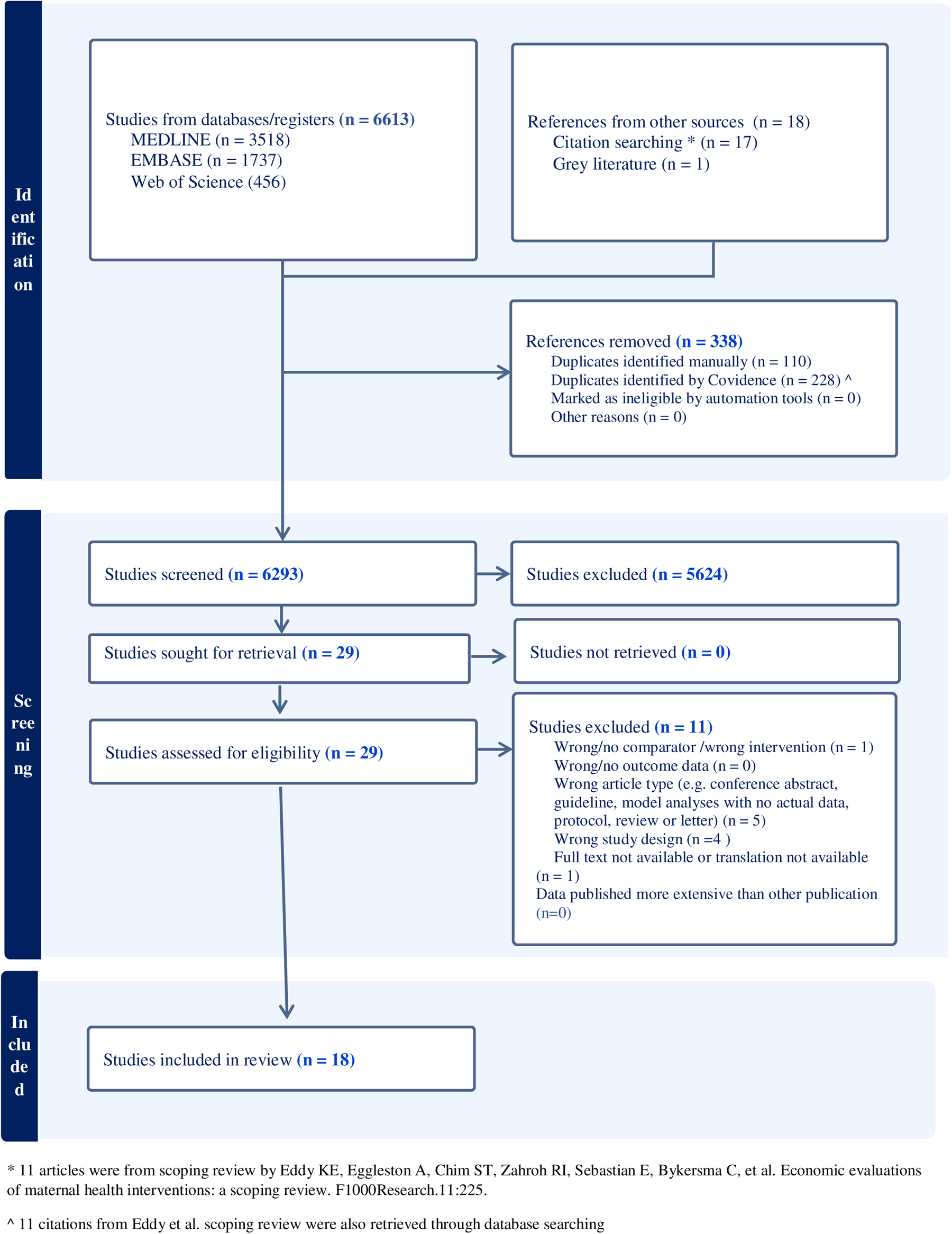
PRISMA flowchart diagram

#### Screening and Extraction

Studies identified by the search were uploaded to the Covidence software (https://www.covidence.org) where de-duplication took place. Three authors (CLA, TL and TP) independently screened potential economic studies against eligibility criteria for inclusion. This was performed firstly by title-abstract and subsequently by full text, conflicts that arose in this process were resolved by discussion. Studies that met the eligibility criteria were included for data extraction, which was performed by two authors independently (CLA, TL). Key characteristics extracted were: bibliographic information (authors, study title, country world bank income level, year of cost estimates, gestational age of intervention, type of economic evaluation, analytic viewpoint (perspective), year of publication, author(s), country of publication, as well as data relating to the comparator, intervention, results, cost-effective (yes/no) and the author conclusions.

#### Assessment of quality

We applied a quality scoring of the selected articles by using the Consensus Health Economic Criteria (CHEC) tool (18) focusing on the methodological quality of economic evaluation. Two reviewers (MEH and EN) assigned a quality score for each criterion, scaled it to a value between 0 and 20 (inclusive) and then divided the maximum possible score by 20, based on set criteria. Finally, the two reviewers compared quality scores and reached a consensus for each study. The studies were then categorised as high (>0.75), moderate (>0.5 and <=0.75) and low (<=0.5).

## Results

### Studies included in the analysis

The search of bibliographic databases yielded 6,293 studies to be screened at title and abstract level following which 29 progressed to be evaluated at the full text level. After full text review 18 studies were eligible for inclusion in this review (Figure 1). Studies that were excluded at full text stage are presented in Suppl. Table 3.

#### Characteristics of studies included

All 18 studies were conducted in high-income countries (HICs): ten in the United States of America (USA), two in the United Kingdom (UK), two in France and one each in Australia, Israel, Switzerland and the Netherlands. The study characteristics are presented in Table 1. The majority of studies (13/18) conducted only one type of economic analysis however a minority (5/18) presented multiple types/combinations in their results. The most common type of economic evaluation conducted were cost-effectiveness analyses (CEA) conducted by nine studies, followed by cost-utility analyses (CUA) in seven studies and cost-benefit analyses (CBA) and cost-consequence analyses (CCA) each conducted in three studies. Studies presented their findings depending on the type of economic evaluation that they had conducted. CEAs and CUAs most frequently presented incremental cost-effectiveness ratios (ICERs), which signify the cost of choosing an intervention over its comparator in terms of the clinical benefits arising as a result (19). CEAs also presented their ICERs in terms of cost per a discrete clinical outcome(s). In this review, studies presented one or both of the following two outcomes: cost per case of EOGBS prevented and cost per EOGBS-associated neonatal death averted. The CUAs included in this review, which used composite outcome measures (e.g., QALYs), were heterogeneous in the outcomes that formed these. Of the three CBAs included, one presented their findings as benefit-cost ratios, the other as net societal benefits, the third presented both. Of the three CCAs, one reported cost-savings per case of neonatal sepsis detected, another compared the average cost per delivery in each of its intervention arms, and the third calculated the extra cost of PCR required to avoid one additional case of EOGBS.

**Table 1.**
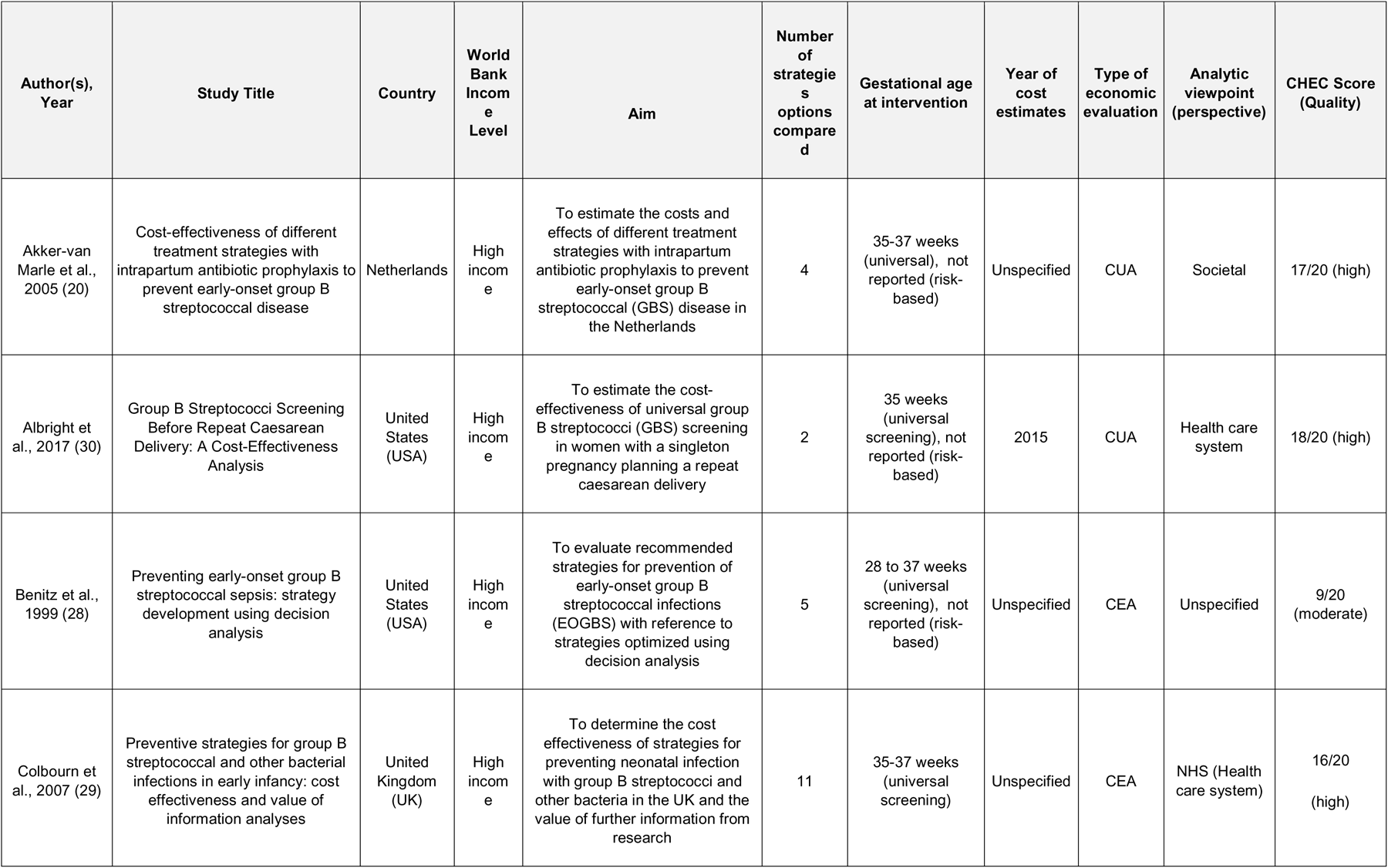
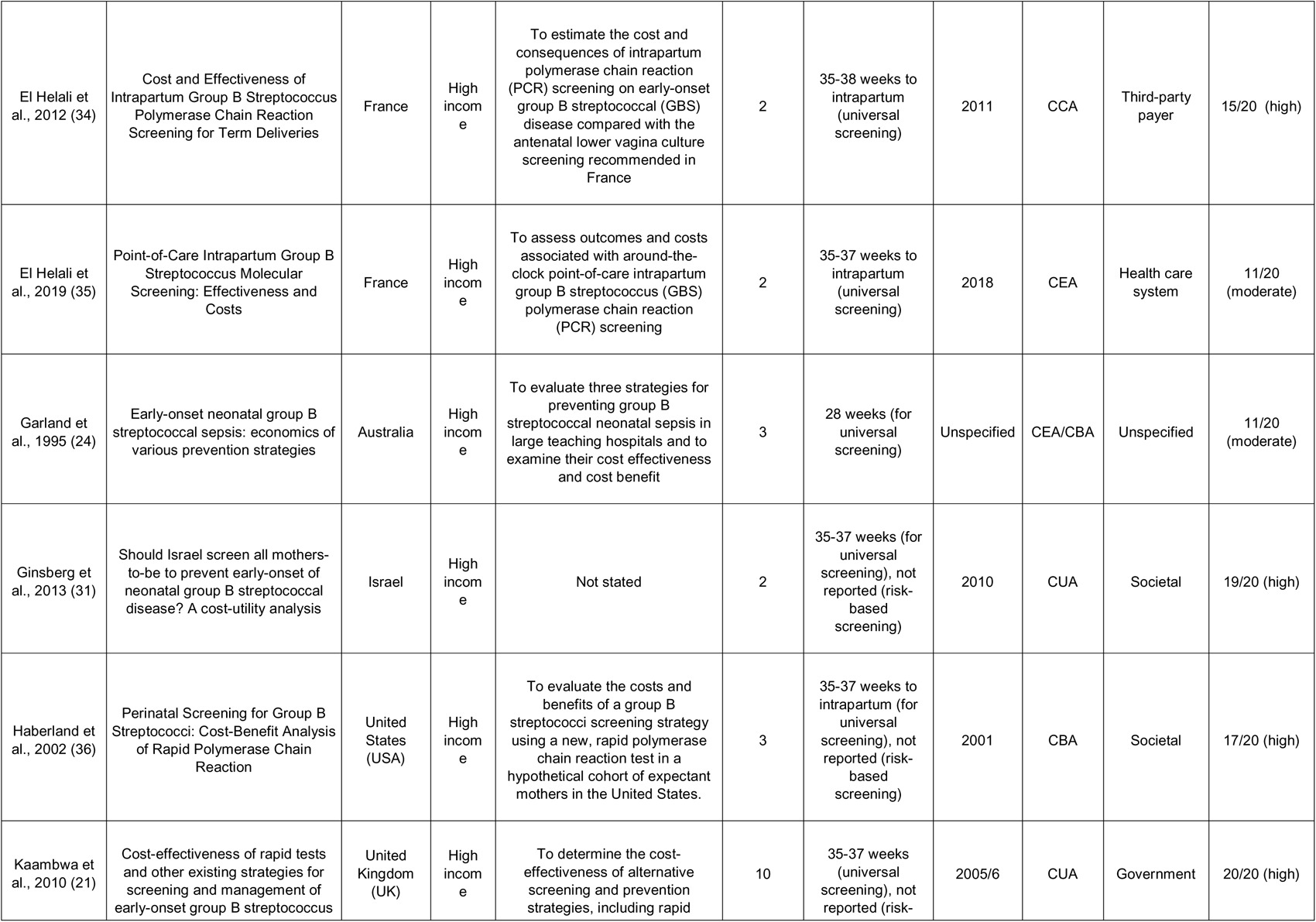
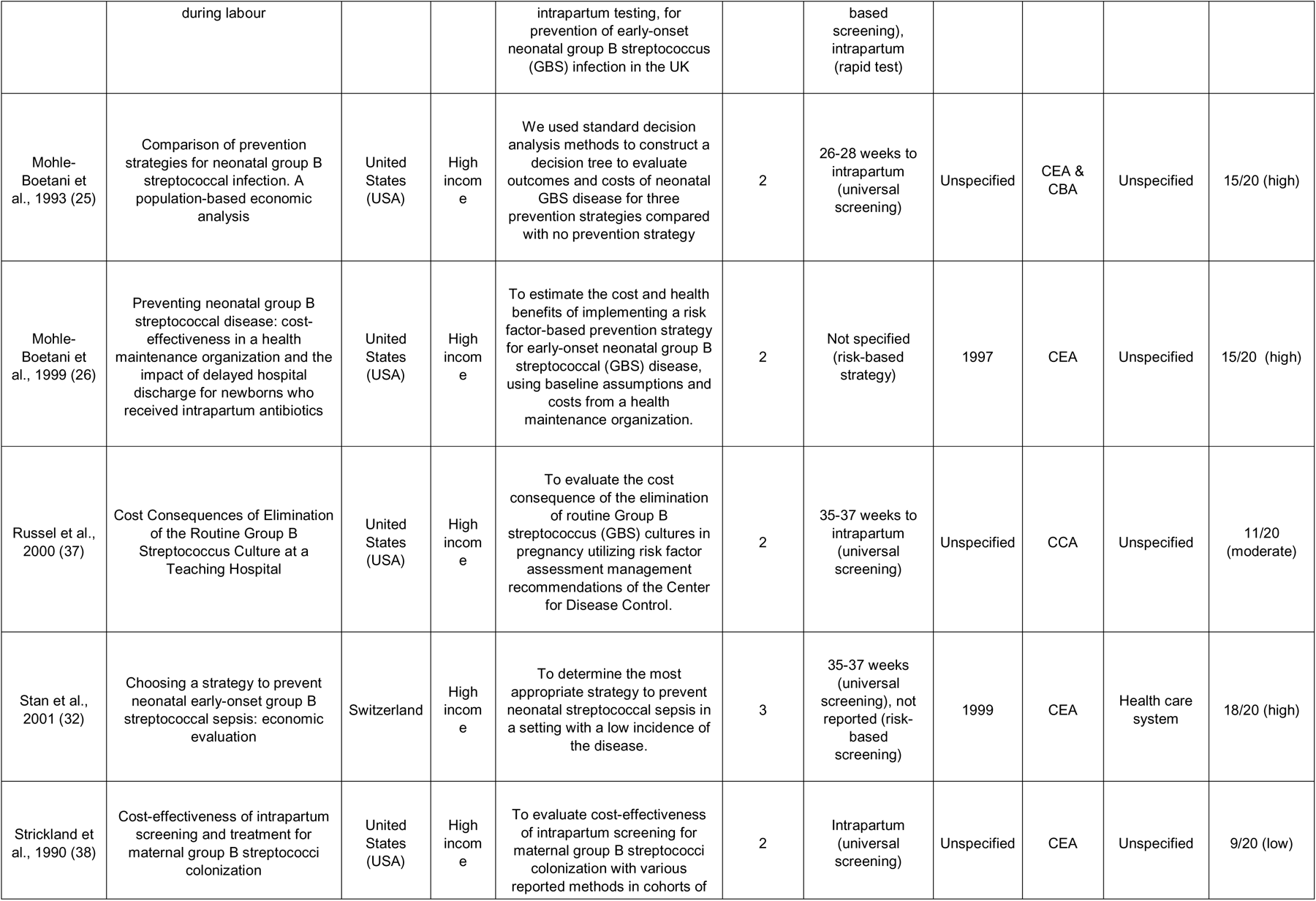
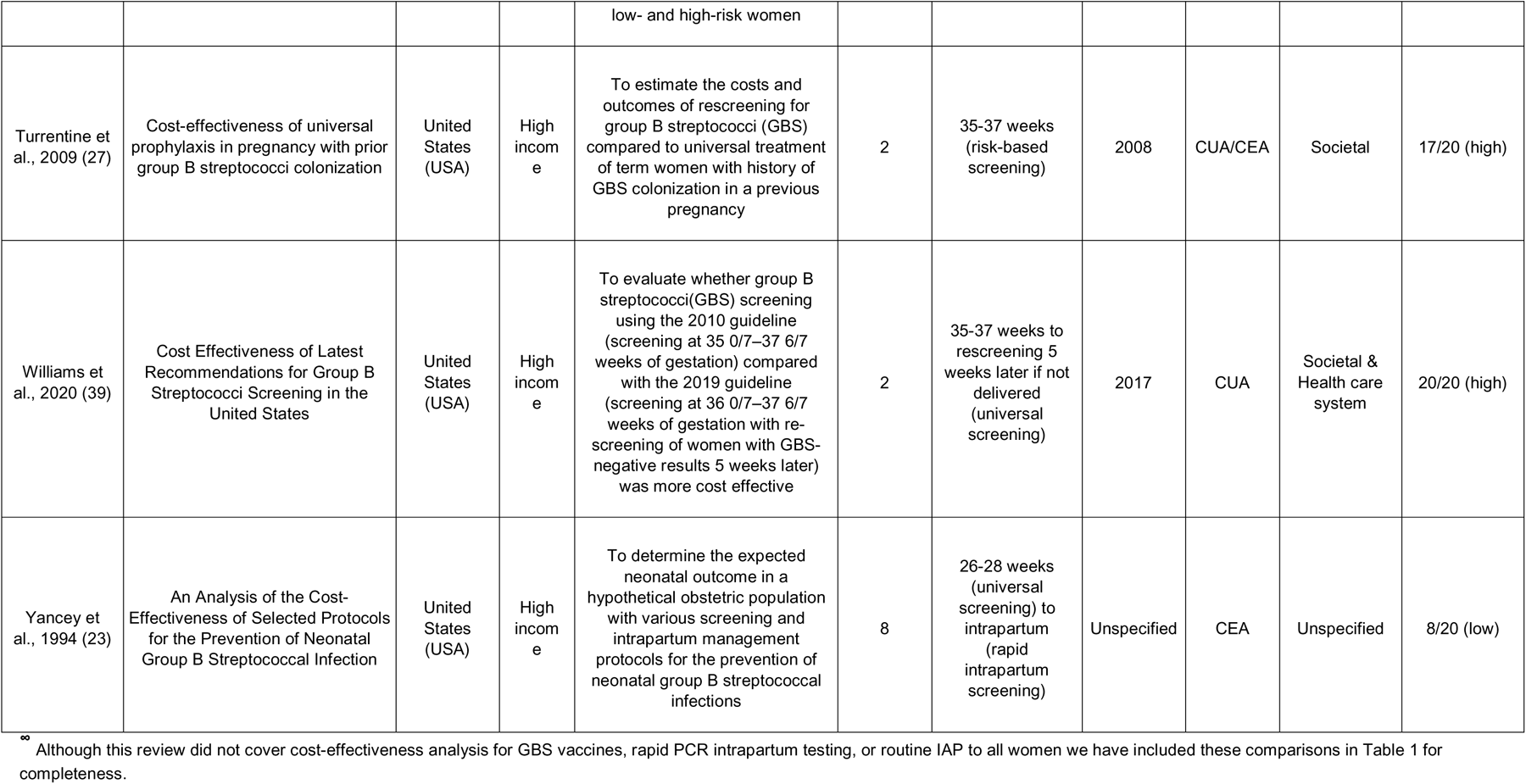
Characteristics of studies included.^∞^.

| Author(s),<br>Year | Study Title | Country | World<br>Bank<br>Income<br>Level | Aim | Number<br>of<br>strategies<br>options<br>compared | Gestational age<br>at intervention | Year of<br>cost<br>estimates | Type of<br>economic<br>evaluation | Analytic<br>viewpoint<br>(perspective) | CHEC Score<br>(Quality) |
| --- | --- | --- | --- | --- | --- | --- | --- | --- | --- | --- |
| Akker-van<br>Marle et al.,<br>2005 (20) | Cost-effectiveness of different treatment strategies with intrapartum antibiotic prophylaxis to prevent early-onset group B streptococcal disease | Netherlands | High income | To estimate the costs and effects of different treatment strategies with intrapartum antibiotic prophylaxis to prevent early-onset group B streptococcal (GBS) disease in the Netherlands | 4 | 35-37 weeks (universal), not reported (risk-based) | Unspecified | CUA | Societal | 17/20 (high) |
| Albright et al., 2017 (30) | Group B Streptococci Screening Before Repeat Caesarean Delivery: A Cost-Effectiveness Analysis | United States (USA) | High income | To estimate the cost-effectiveness of universal group B streptococci (GBS) screening in women with a singleton pregnancy planning a repeat caesarean delivery | 2 | 35 weeks (universal screening), not reported (risk-based) | 2015 | CUA | Health care system | 18/20 (high) |
| Benitz et al., 1999 (28) | Preventing early-onset group B streptococcal sepsis: strategy development using decision analysis | United States (USA) | High income | To evaluate recommended strategies for prevention of early-onset group B streptococcal infections (EOGBS) with reference to strategies optimized using decision analysis | 5 | 28 to 37 weeks (universal screening), not reported (risk-based) | Unspecified | CEA | Unspecified | 9/20 (moderate) |
| Colbourn et al., 2007 (29) | Preventive strategies for group B streptococcal and other bacterial infections in early infancy: cost effectiveness and value of information analyses | United Kingdom (UK) | High income | To determine the cost effectiveness of strategies for preventing neonatal infection with group B streptococci and other bacteria in the UK and the value of further information from research | 11 | 35-37 weeks (universal screening) | Unspecified | CEA | NHS (Health care system) | 16/20 (high) |
| El Helali et al., 2012 (34) | Cost and Effectiveness of Intrapartum Group B Streptococcus Polymerase Chain Reaction Screening for Term Deliveries | France | High income | To estimate the cost and consequences of intrapartum polymerase chain reaction (PCR) screening on early-onset group B streptococcal (GBS) disease compared with the antenatal lower vagina culture screening recommended in France | 2 | 35-38 weeks to intrapartum (universal screening) | 2011 | CCA | Third-party payer | 15/20 (high) |
| El Helali et al., 2019 (35) | Point-of-Care Intrapartum Group B Streptococcus Molecular Screening: Effectiveness and Costs | France | High income | To assess outcomes and costs associated with around-the-clock point-of-care intrapartum group B streptococcus (GBS) polymerase chain reaction (PCR) screening | 2 | 35-37 weeks to intrapartum (universal screening) | 2018 | CEA | Health care system | 11/20 (moderate) |
| Garland et al., 1995 (24) | Early-onset neonatal group B streptococcal sepsis: economics of various prevention strategies | Australia | High income | To evaluate three strategies for preventing group B streptococcal neonatal sepsis in large teaching hospitals and to examine their cost effectiveness and cost benefit | 3 | 28 weeks (for universal screening) | Unspecified | CEA/CBA | Unspecified | 11/20 (moderate) |
| Ginsberg et al., 2013 (31) | Should Israel screen all mothers-to-be to prevent early-onset of neonatal group B streptococcal disease? A cost-utility analysis | Israel | High income | Not stated | 2 | 35-37 weeks (for universal screening), not reported (risk-based screening) | 2010 | CUA | Societal | 19/20 (high) |
| Haberland et al., 2002 (36) | Perinatal Screening for Group B Streptococci: Cost-Benefit Analysis of Rapid Polymerase Chain Reaction | United States (USA) | High income | To evaluate the costs and benefits of a group B streptococci screening strategy using a new, rapid polymerase chain reaction test in a hypothetical cohort of expectant mothers in the United States. | 3 | 35-37 weeks to intrapartum (for universal screening), not reported (risk-based screening) | 2001 | CBA | Societal | 17/20 (high) |
| Kaambwa et al., 2010 (21) | Cost-effectiveness of rapid tests and other existing strategies for screening and management of early-onset group B streptococcus | United Kingdom (UK) | High income | To determine the cost-effectiveness of alternative screening and prevention strategies, including rapid | 10 | 35-37 weeks (universal screening), not reported (risk- | 2005/6 | CUA | Government | 20/20 (high) |
|  | during labour |  |  | intrapartum testing, for prevention of early-onset neonatal group B streptococcus (GBS) infection in the UK |  | based screening), intrapartum (rapid test) |  |  |  |  |
| Mohle-Boetani et al., 1993 (25) | Comparison of prevention strategies for neonatal group B streptococcal infection. A population-based economic analysis | United States (USA) | High income | We used standard decision analysis methods to construct a decision tree to evaluate outcomes and costs of neonatal GBS disease for three prevention strategies compared with no prevention strategy | 2 | 26-28 weeks to intrapartum (universal screening) | Unspecified | CEA & CBA | Unspecified | 15/20 (high) |
| Mohle-Boetani et al., 1999 (26) | Preventing neonatal group B streptococcal disease: cost-effectiveness in a health maintenance organization and the impact of delayed hospital discharge for newborns who received intrapartum antibiotics | United States (USA) | High income | To estimate the cost and health benefits of implementing a risk factor-based prevention strategy for early-onset neonatal group B streptococcal (GBS) disease, using baseline assumptions and costs from a health maintenance organization. | 2 | Not specified (risk-based strategy) | 1997 | CEA | Unspecified | 15/20 (high) |
| Russel et al., 2000 (37) | Cost Consequences of Elimination of the Routine Group B Streptococcus Culture at a Teaching Hospital | United States (USA) | High income | To evaluate the cost consequence of the elimination of routine Group B streptococcus (GBS) cultures in pregnancy utilizing risk factor assessment management recommendations of the Center for Disease Control. | 2 | 35-37 weeks to intrapartum (universal screening) | Unspecified | CCA | Unspecified | 11/20 (moderate) |
| Stan et al., 2001 (32) | Choosing a strategy to prevent neonatal early-onset group B streptococcal sepsis: economic evaluation | Switzerland | High income | To determine the most appropriate strategy to prevent neonatal streptococcal sepsis in a setting with a low incidence of the disease. | 3 | 35-37 weeks (universal screening), not reported (risk-based screening) | 1999 | CEA | Health care system | 18/20 (high) |
| Strickland et al., 1990 (38) | Cost-effectiveness of intrapartum screening and treatment for maternal group B streptococci colonization | United States (USA) | High income | To evaluate cost-effectiveness of intrapartum screening for maternal group B streptococci colonization with various reported methods in cohorts of low- and high-risk women | 2 | Intrapartum (universal screening) | Unspecified | CEA | Unspecified | 9/20 (low) |
| Turrentine et al., 2009 (27) | Cost-effectiveness of universal prophylaxis in pregnancy with prior group B streptococci colonization | United States (USA) | High income | To estimate the costs and outcomes of rescreening for group B streptococci (GBS) compared to universal treatment of term women with history of GBS colonization in a previous pregnancy | 2 | 35-37 weeks (risk-based screening) | 2008 | CUA/CEA | Societal | 17/20 (high) |
| Williams et al., 2020 (39) | Cost Effectiveness of Latest Recommendations for Group B Streptococci Screening in the United States | United States (USA) | High income | To evaluate whether group B streptococci(GBS) screening using the 2010 guideline (screening at 35 0/7–37 6/7 weeks of gestation) compared with the 2019 guideline (screening at 36 0/7–37 6/7 weeks of gestation with re-screening of women with GBS-negative results 5 weeks later) was more cost effective | 2 | 35-37 weeks to rescreening 5 weeks later if not delivered (universal screening) | 2017 | CUA | Societal & Health care system | 20/20 (high) |
| Yancey et al., 1994 (23) | An Analysis of the Cost-Effectiveness of Selected Protocols for the Prevention of Neonatal Group B Streptococcal Infection | United States (USA) | High income | To determine the expected neonatal outcome in a hypothetical obstetric population with various screening and intrapartum management protocols for the prevention of neonatal group B streptococcal infections | 8 | 26-28 weeks (universal screening) to intrapartum (rapid intrapartum screening) | Unspecified | CEA | Unspecified | 8/20 (low) |

<sup>oo</sup> Although this review did not cover cost-effectiveness analysis for GBS vaccines, rapid PCR intrapartum testing, or routine IAP to all women we have included these comparisons in Table 1 for completeness.

Given the heterogeneity of results, data were synthesised and analysed narratively.

#### Quality appraisal

According to the CHEC tool and quality scoring, this review consisted of 12 high quality, four moderate quality and two low quality studies (Suppl. Table 4 and 5 show the assessment for each study).

#### Technical characteristics of the studies

The majority of studies did not specify an analytic viewpoint/perspective employed when conducting economic analyses, considering the perspective of the economic evaluation is crucial, as it determines which costs and effects should be included in the study. Few authors explicitly stated the perspective of their articles. However, after reviewing the articles, the perspective of the studies could often be understood even if it was not explicitly mentioned. In studies a societal perspective was most commonly adopted (4), followed by health care system (4), societal & health care system (1), third-party payer (1) and governmental (1).

All studies identified the costs incurred based on the different intervention alternatives. Most of the studies measured outcomes using natural units, while a few used quality-adjusted life years (QALYs) as the outcome measurement. Several studies measured multiple outcomes of same intervention. Discounting has been applied in few studies, though justification of considering specific discounting rate was not mentioned properly. Ten studies calculated the incremental cost effectiveness ratio (ICER).

The thresholds by which studies made determinations as to whether an intervention was cost-effective were markedly heterogenous. There were several instances in which the intervention was found to be clinically superior and cost-saving and were therefore deemed dominant thus not requiring consideration against a cost-effectiveness threshold. When this was not the case, the majority of authors determined an intervention’s cost-effectiveness at their own discretion. Other studies were unable to make a final determination of an intervention’s cost-effectiveness. The cost-effectiveness threshold employed by each study is specified in Table 2. Several studies performed sensitivity analysis to assess the robustness of the results to changes in assumptions and values of inputs. No study discussed the generalisability of the result in the context of lower resource settings.

**Table 2.**
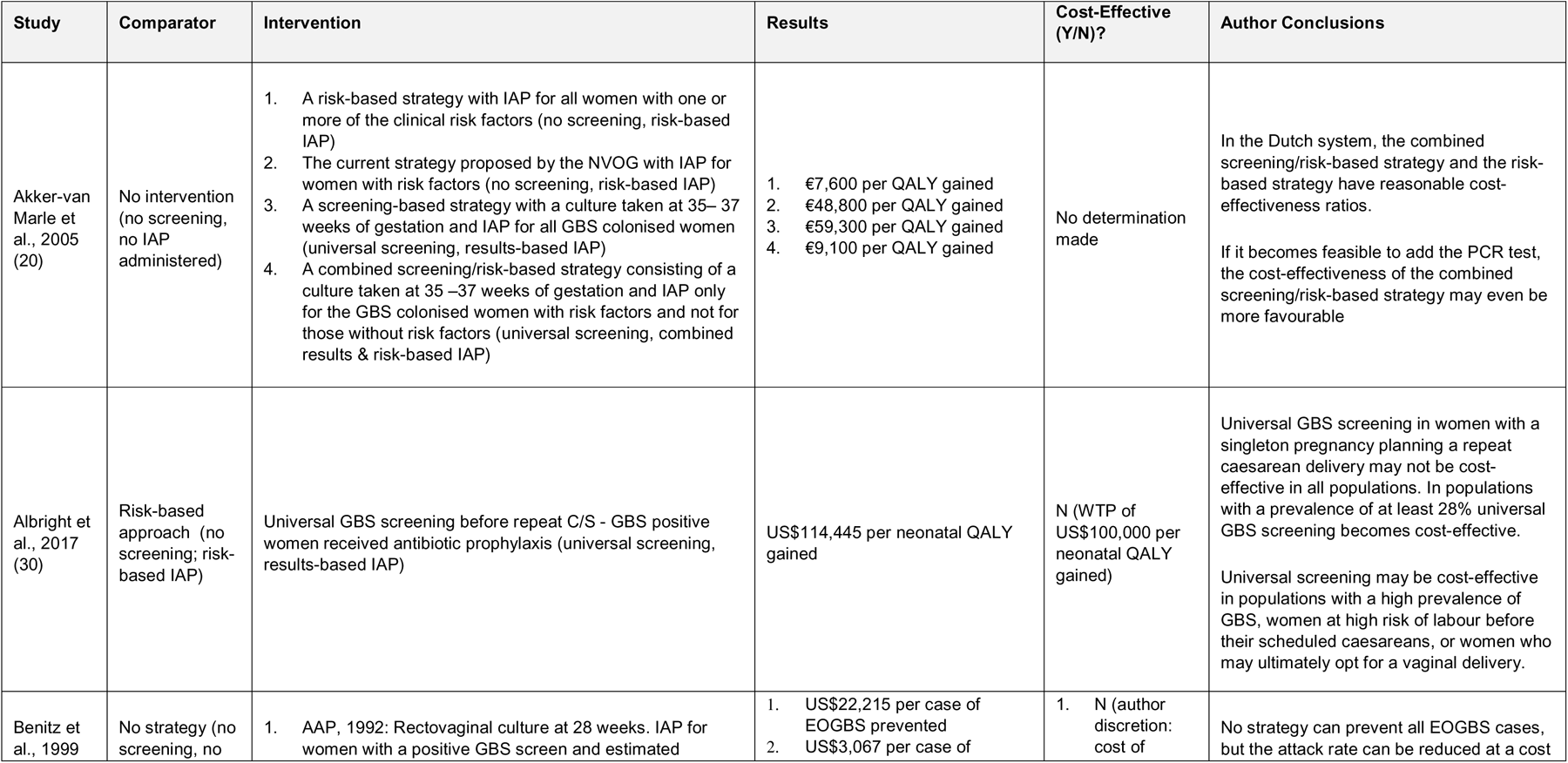
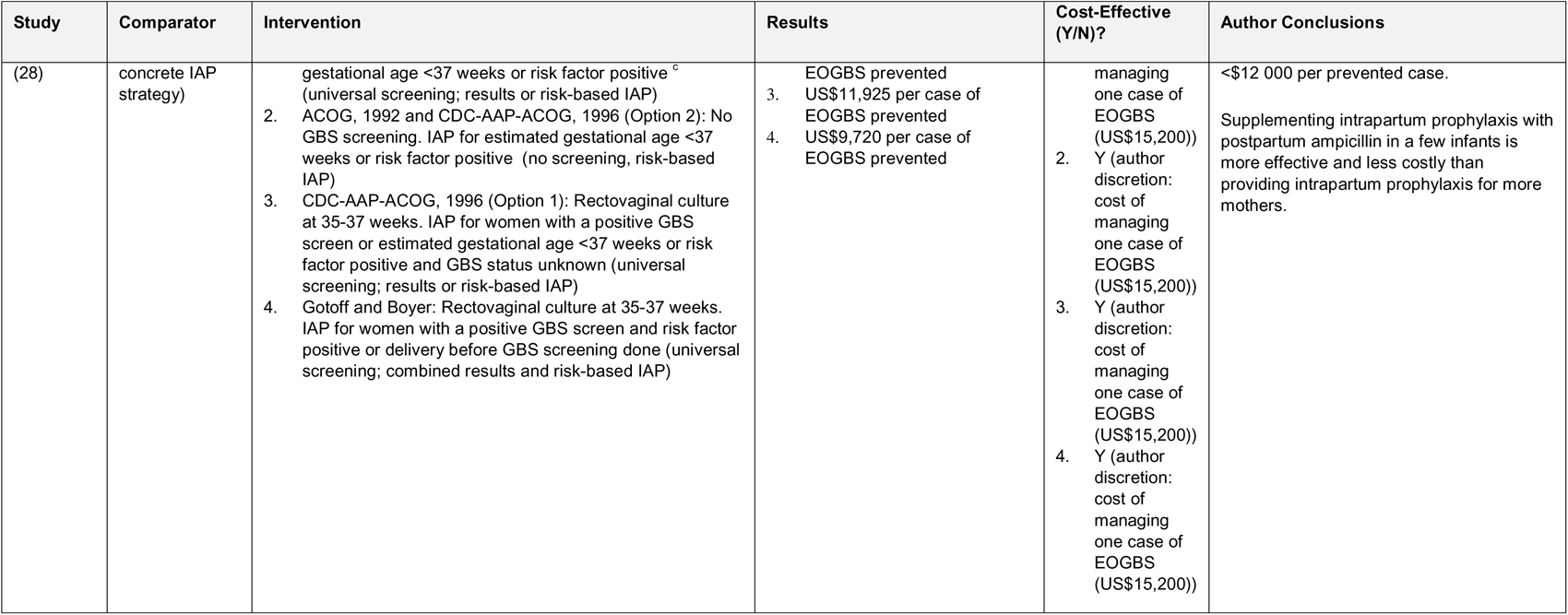
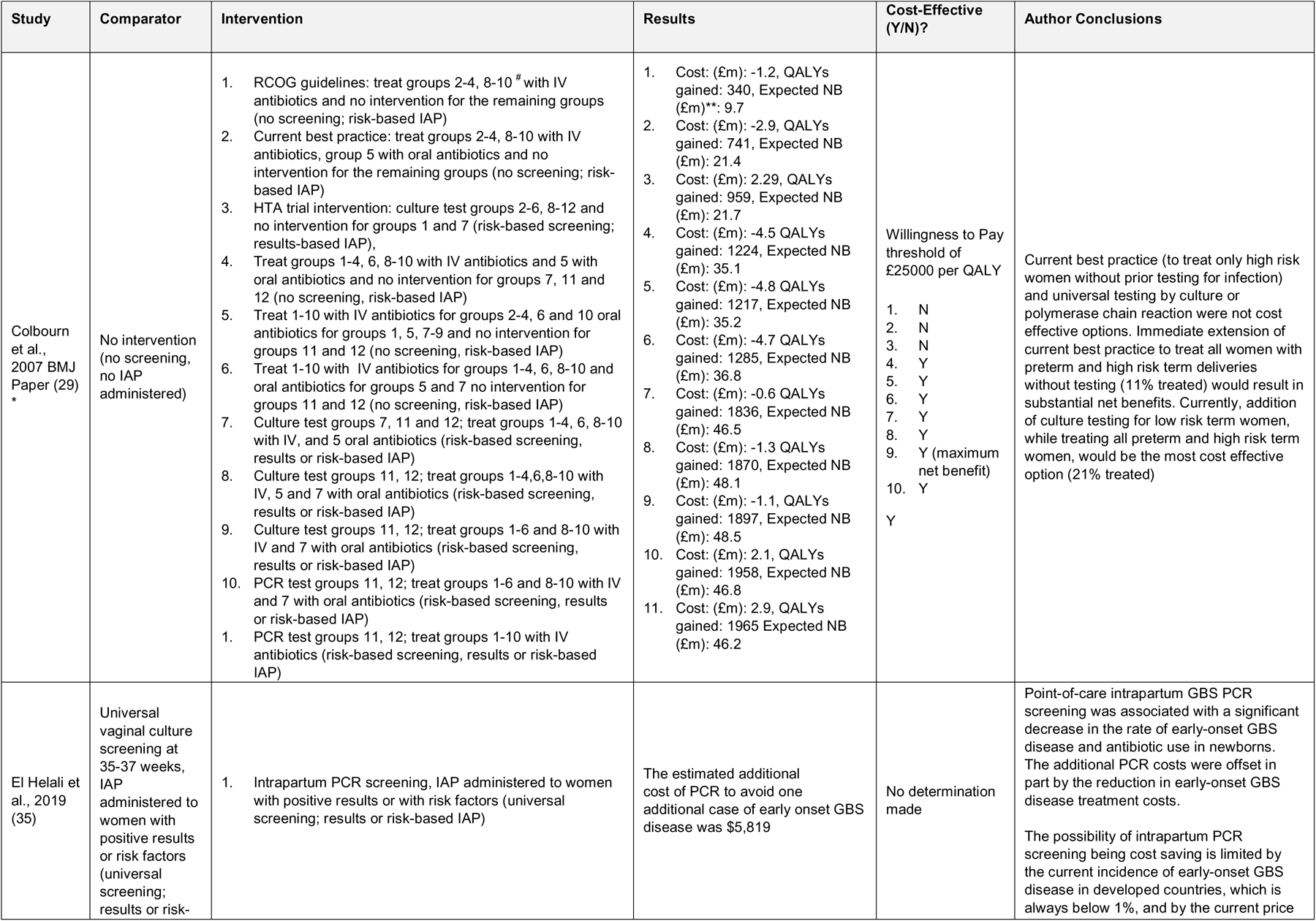
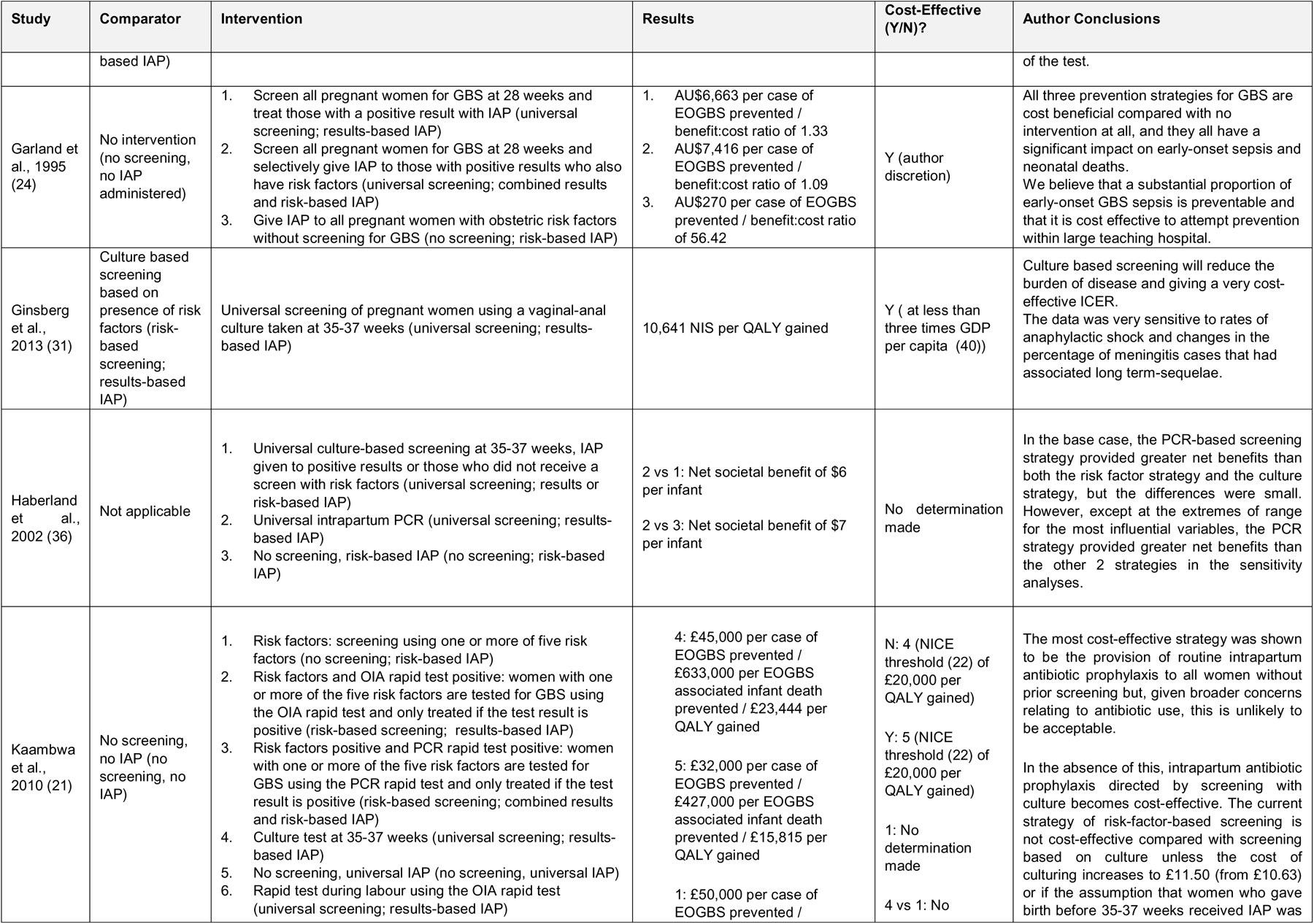
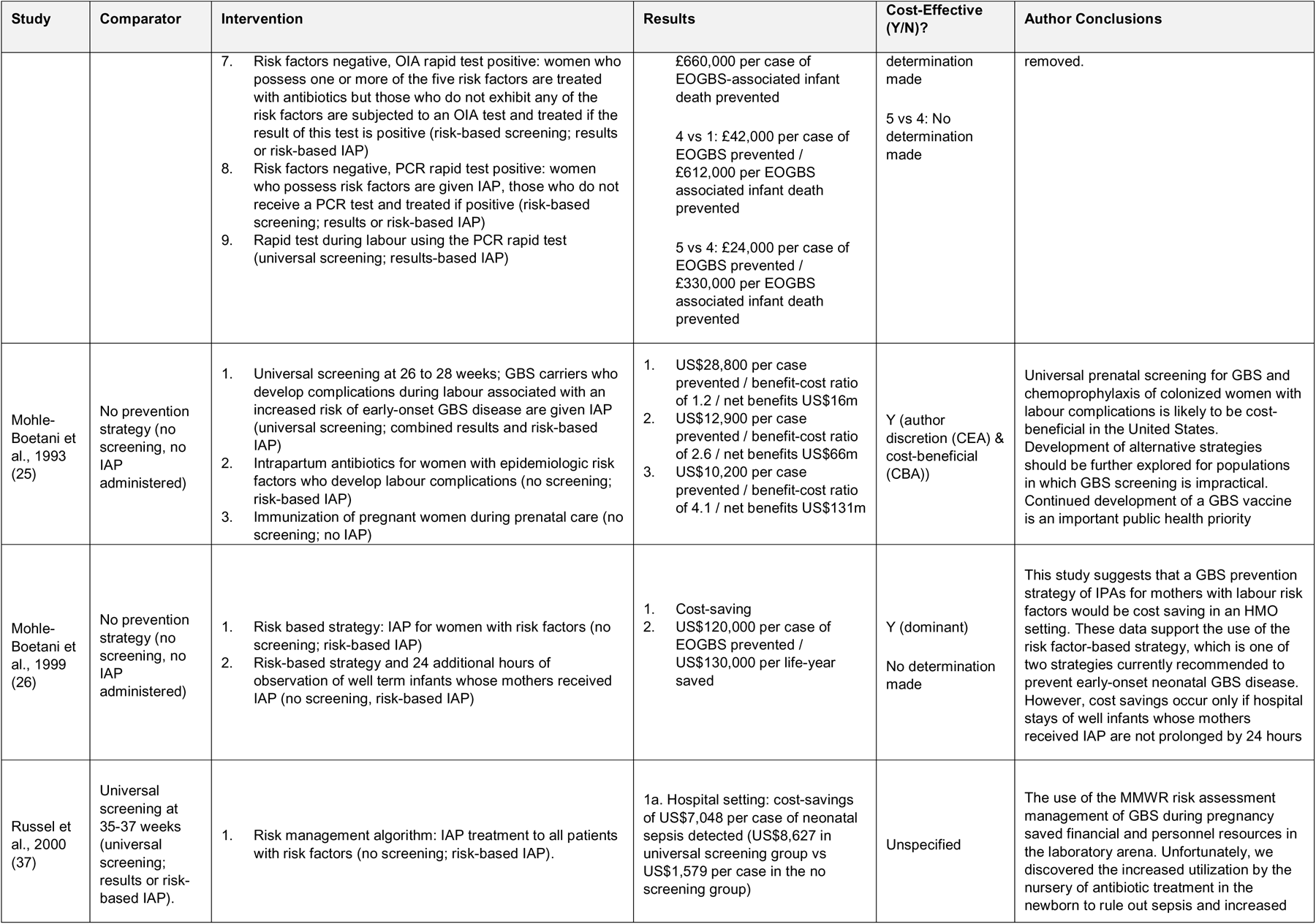
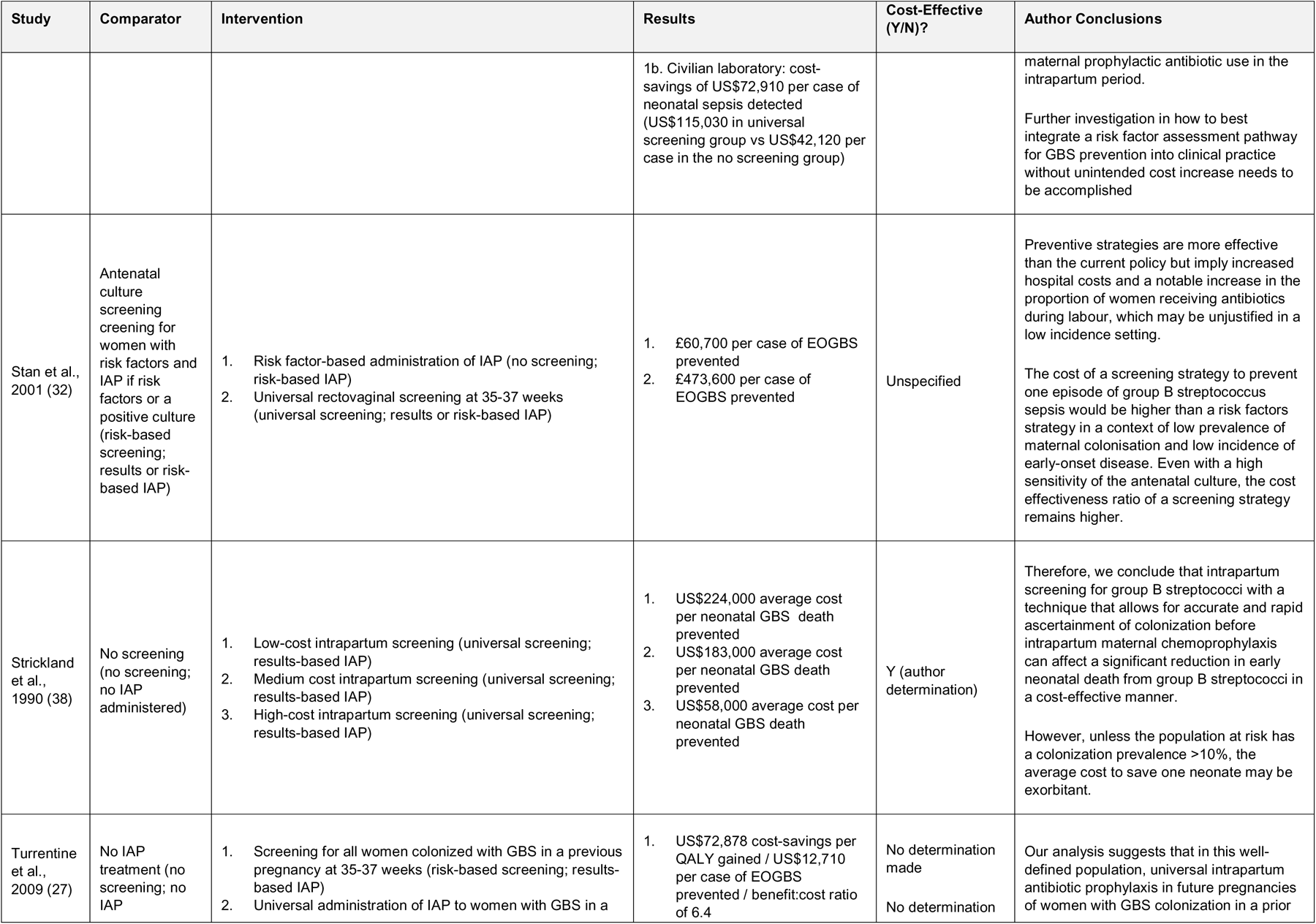
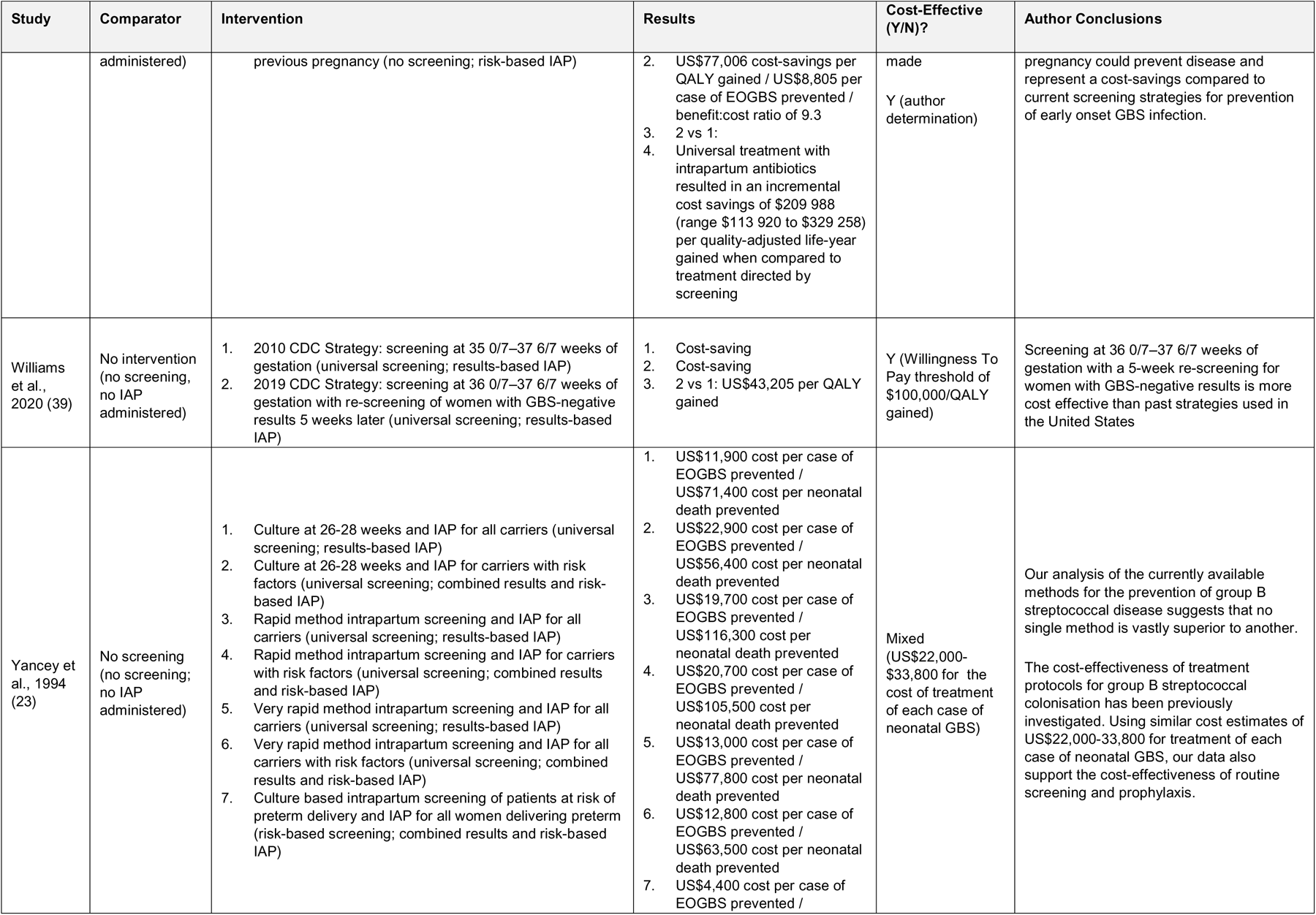
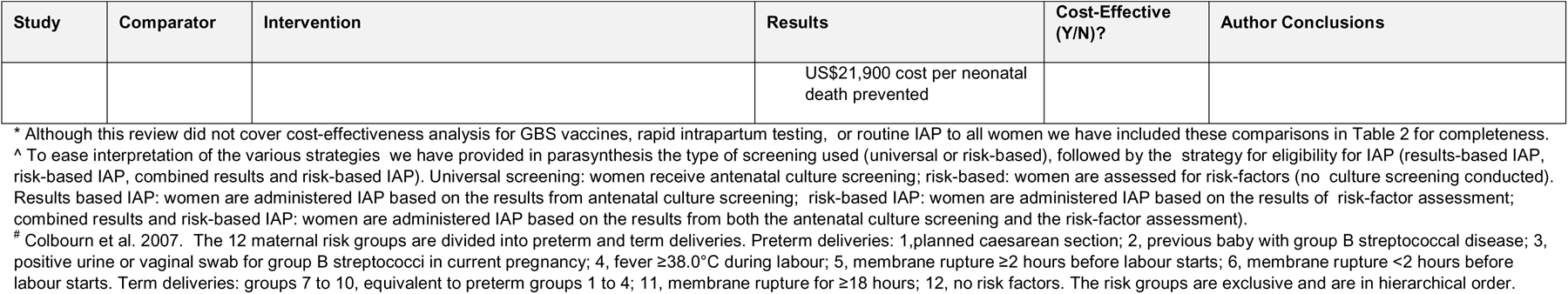
Cost-effectiveness findings of studies included.^∧^.

#### Cost-Effectiveness Findings

The full cost-effectiveness findings for each of the studies included in this review are outlined in Table 2. Due to the large number of strategies for which comparisons were conducted (n=58), the results section is presented by any vs no strategy: (1) universal strategy (with IAP provided to women with positive cultures) vs no strategy; (2) risk-based approach (no cultures conducted and IAP given to women identified with risk-factors for EOGBS) vs no strategy; (3) combined strategy vs no strategy; and three different strategies compared to each other. The latter comparison encompasses (4) combined (any combination of universal and risk-based approach) vs no strategy, (5) universal vs risk-based strategies, (6) universal vs combined strategies, (7) risk-based vs combined strategies. Although this review did not include cost-effectiveness analyses for GBS vaccines, rapid PCR intrapartum testing, or routine IAP to all women, we have included these comparisons in Table 2 for completeness.

#### Universal screening vs no screening

Universal screening (with IAP provided to women who screened positive) vs no screening was compared in four studies (three high quality, one low quality) in which five ICERs were calculated. Two studies conducted sensitivity analyses investigating the impact of maternal GBS prevalence on the cost-effectiveness of universal screening; in one of these there was no relationship identified, in the other, cost-effectiveness was optimised at higher rates of maternal GBS prevalence.

ME Van den (2005) (20) investigated the cost-effectiveness of a universal screening strategy using a culture at 35-37 weeks with IAP administered to all women colonised with GBS compared to no strategy. The proposed screening strategy was found to cost €59,300 per QALY gained however no final determination of its cost-effectiveness was made. Another study conducted in 2010 (21) compared to no intervention, this strategy cost £45,000 per case of EOGBS prevented or £23,444 per QALY gained. The author determined that this would not be cost-effective when evaluated against the NICE threshold of £20,000 per QALY gained (22).

Yancey MK (1994) (23) compared universal screening with a culture at 26-28 weeks with subsequent IAP administered to those who test positive against a strategy of no intervention. In this instance, the universal screening strategy would cost US $11,900 per case of EOGBS prevented. When evaluated against a threshold of US $22,000-$33,800, which the author estimates as the cost for treatment of each case of EOGBS, this strategy of universal screening was deemed cost-effective.

#### Risk-based approach vs no screening

A risk-based approach vs no screening and no IAP (no intervention) was the most common comparison identified by this review and was represented in seven studies (five high quality, two moderate quality). Compared to no screening strategy, two studies found risk-based approaches to be cost-effective, none found it to be less cost-effective, while four did not make a final determination of whether it was more or less cost-effective. Three of these studies conducted sensitivity analyses investigating the impact of the prevalence of maternal GBS colonization on cost-effectiveness; one study found no relationship between maternal GBS prevalence and cost-effectiveness while two studies identified that cost-effectiveness of risk-based screening increased when maternal GBS prevalence was higher.

ME Van den (2005) (20) compared no intervention to a risk-based strategy with IAP to women with one or more clinical risk-factors (preterm birth (<37 weeks), prelabour rupture of membranes (>18 hours)). Compared to no screening, the risk-based strategy cost €7,600 per QALY gained. The author did not make a final determination as to whether this was cost-effective but described the risk-based strategy as having a “reasonable cost-effectiveness ratio”.

Kaambwa B (2010) (21) compared risk-based administration of IAP, determined by the presence of at least one risk factor (not reported), compared to no intervention. Switching to this strategy would cost £50,000 per case of EOGBS prevented. The author did not make an outright determination as to whether this was cost-effective.

Garland SM (1995) (24) conducted a study in Australia to compared giving IAP to all pregnant women with risk factors (preterm labour <37 weeks, prolonged rupture of membranes >12 hours or maternal sepsis) compared to no intervention. Implementing this strategy costs AU$270 per case of EOGBS prevented when compared to no intervention; as per the author’s discretion this was deemed cost-effective.

Mohle-Boetani JC (1993) (25) compared no intervention to IAP administration to women with epidemiological risk factors, specifically if the mother was a teenager or Black, who had also developed labour complications including fever (>37.5°), preterm labour (<37 weeks) or prolonged rupture of membranes (>12 hours). Implementation of this risk-based strategy was found to cost US$12,900 per case of EOGBS prevented. CBA demonstrated the benefit-cost ratio of this strategy to be 2.6 compared to no intervention, with annual savings of approximately $66m. Sensitivity analysis demonstrated that the rate of EOGBS at which this strategy became cost-saving was 0.65 per 1000 live births.

Mohle-Boetani JC (1999) (26) evaluated two risk-based strategies compared to no intervention. Both strategies consisted of providing IAP to women with certain risk factors, however one included 24 additional hours of observation for the well term infants of mothers who had received IAP. Compared to no intervention, the risk-based strategy of IAP administration without additional observation of infants was cost-saving and therefore dominant. The risk-based strategy of IAP administration with an additional 24 hours of infant observation bore a net cost of US$120,000 per case of EOGBS prevented or US$130,000 per life-year saved. The author did not make a final determination as to whether this second strategy was cost-effective.

Turrentine MA (2009) (27) evaluated the cost-effectiveness of administering IAP to all pregnant women with a history of GBS colonisation in a previous pregnancy compared to no intervention. Implementing this strategy would cost an additional US$8,805 per case of EOGBS prevented compared to no intervention. The author did not make a determination as to whether this is cost-effective.

#### Combined screening vs no screening

Six studies reported on a combined screening approach as compared to no screening. These studies were largely heterogenous in their combined strategy but could be grouped into two broad strategies: (1) antenatal culture screening with IAP provided only to women with both GBS colonisation and risk factors (20, 23–25, 28) (2) women with specific risk-factors have culture screening, while women with other risk-factors or absence of risk-factors are not screened using culture; with IAP eligibility based on combined risk-factor and GBS colonisation results (29).

#### Antenatal culture screening with IAP only to women with both GBS colonisation and risk-factors vs no screening

Akker-van Marle et al. (2005) reported the combined strategy resulted in a cost of 9,100 euro per QALY gained, however the authors did not specify a cost-effectiveness threshold or make a determination on cost-effectiveness (20). Benitz et al. (1999) found that the combined strategy was not cost-effective when screening at 28 weeks (USD $22,215 per case of EOGBS prevented) but was cost-effective when screening at 35-37 weeks gestation (USD $ 15,200 per case of EOGBS prevented) (28). Garland et al. (2005) found a combined strategy with antenatal culture screening at 28 weeks to be cost-effective (AU$7,416 per case of EOGBS prevented) (24). Mohle-Boetani et al. 1993 found that a combined strategy using screening at 26-28 weeks was cost-effective (USD $28,800 per case of EOGBS prevented) (25). Yancey et al. 1994 did not make a determination on cost-effectiveness but reported that the cost was between USD $4,400 and USD $22,900 depending on the risk-factors used for the risk-based approach (23).

#### Women with specific risk-factors have culture screening with IAP eligibility based on risk-factor and culture results

Colbourn et al. 2007 (29) reported on combined screening approaches where women divided into 12 different maternal risk groups using a risk-based approach. Some maternal risk groups had culture screening, while other maternal risk groups did not. IAP eligibility was based on both risk-factors and culture screening results. The combined strategies were found to be cost-effective with a threshold of GBP 25,000 per QALY gained.

#### Universal screening vs risk-based approach

Albright CM (2017) (30) compared the cost-effectiveness of universal GBS screening to a risk-based approach for women presenting for repeat caesareans. The cost of the implementing the universal screening program instead of the risk-based approach was US$114,445 per neonatal QALY gained. When evaluated against a willingness-to-pay threshold (WTP) of US$100,000 per neonatal QALY gained, the authors determined that the universal screening program was not cost-effective. Sensitivity analysis demonstrated that in settings where maternal GBS colonisation has a prevalence of at least 28%, universal screening would become cost-effective compared to a risk-based approach.

#### Universal screening vs combined screening

Ginsberg et al. 2013 reported on universal screening as compared to a combined strategy of culture based screening only to women with risk-factors (31). This strategy was found to be cost-effective when using a threshold of less than three times GDP per capita. Stan et al. 2001 reported on universal screening as compared to a combined strategy where only women with risk-factors were screened and IAP eligibility was based on either risk-factors or a positive culture (32). The authors did not determine cost-effectiveness but found that the cost of implementing universal instead of combined screening was GBP 473,600 per case of EOGBS prevented.

#### Risk-based approach vs combined screening

Stan et al. 2001 compared a risk-based approach to a combined strategy of antenatal culture screening to women with risk-factors with IAP based on risk-factors or positive culture (32). The cost of a risk-based approach instead of a combined screening strategy cost was GBP 60,700 per case of EOGBS prevented. The author did not determine whether this was cost-effective.

## Discussion

### Main findings

This study demonstrated several instances where the implementation of a GBS screening strategy for pregnant women to reduce the incidence of newborn EOGBS would be both clinically and economically advantageous compared to no intervention.

Seventeen studies compared any strategy (4 universal screening, 7 risk-based approach, 6 combined screening) to no screening strategy, several studies presented multiple ICERs. In total there were seven studies in which the implementation of at least one GBS prevention strategy (one using universal screening, two using risk-based approach, four using combined screening) was found to be cost-effective when evaluated against no intervention, one study in which the proposed screening strategy was not cost-effective (universal) and seven instances (one using universal screening, four using risk-based approach, two using combined screening) where final cost-effectiveness was not determined.

Comparisons between specific screening strategies were limited. Only one study compared universal screening compared to a risk-based strategy in women with repeat caesarean section; this study ultimately concluded that opting for a universal screening was not cost-effective. Similarly, of the two studies that compared universal and combined screening, only one identified that opting for universal screening was cost-effective, the other made no final determination. In the one study that compared a risk-based approach and combined screening, no conclusion as to which was more cost-effective was provided. Greater evidence comparing different screening strategies will be required to better understand which screening strategies are most economically advantageous and in which contexts.

### Strengths and limitations

To our knowledge, this review is the first to compile existing cost-effectiveness evidence for maternal GBS screening strategies. One strength of this review was the range and diversity of screening and prevention strategies that it included and facilitated comparisons between. In collating such a broad body of evidence and diversity of screening strategies, we were able to elucidate common determinants of cost-effectiveness between studies. These factors, such as higher prevalence of maternal GBS colonisation, could in the future help assess the suitability of proposed screening strategies in various settings.

There are several limitations of this review. Firstly, as all included studies were conducted in HICs, the applicability of this review’s findings to LMICs will be limited as costs, resource availability and disease prevalence will differ greatly in these settings. Typically, the higher a country’s income level the more likely they are to opt for an IAP administration strategy (13). Despite this, the current reality is that low-and middle-income countries (LMICs) are burdened with higher prevalence of GBS colonisation and subsequently experience higher rates of greater morbidity and mortality attributed associated with EOGBS (33). Additionally, there was marked heterogeneity in how cost-effectiveness was determined, both in terms of which clinical outcomes were chosen and in how composite outcomes (QALYs) were derived. The heterogeneity of cost-effectiveness thresholds and the predominance of author made determinations of interventions’ cost-effectiveness impaired a statistical synthesis.

### Clinical significance

This review identified several instances in which cost-effectiveness evidence can be incorporated into decision making processes surrounding the selection and implementation of a GBS screening strategy. Firstly, our review demonstrated that in several instances any screening strategy was cost-effective compared to no strategy at all. Concurrently, although universal screening for GBS has been shown be more effective in reducing the incidence of EOGBS compared to other strategies, our review illustrates that there is limited cost-effectiveness evidence supporting its economic superiority; only one study demonstrating it to be more cost-effective than another type of screening (12). This finding indicates the need for further investigation into determining whether universal screening is a cost-effective way to screen women for GBS compared to other strategies. Risk-based screening as compared to no screening also reduces EOGBS incidence and may be a feasible strategy which was also shown to be cost-effective in two studies. There is a need to explore further which combined strategies are most efficacious as combined strategies were shown to be cost-effective in four instances. The recurrent finding that intervention cost-effectiveness increased in settings with higher prevalence of maternal GBS colonisation should be considered when implementing GBS prevention programs.

## Conclusion

This review demonstrates that the implementation of any screening strategy compared to none is likely to be cost-effective in high-income settings. There were no studies from LMICs limiting the applicability of the findings. In the formulation and implementation of any GBS screening strategy, cost-effectiveness should always be evaluated in the context of other factors such as efficacy, resources and acceptability.

## Data Availability

All data produced in the present work are contained in the manuscript

